# Subjective experience of women with childbirth in hospitals after COVID-19 outbreak: Insights from Babies Born Better survey

**DOI:** 10.1101/2024.12.12.24318966

**Authors:** Katarína Greškovičová, Mariana Němcová, Tereza Šiková

## Abstract

After COVID-19 outbreak, there has been changes in maternity care suggesting that childbirth experience was also change. This study thus investigates the impact of the COVID-19 pandemic on childbirth experiences in Slovak hospitals, focusing on women’s perspectives to childbirth experience. Utilizing data from the Babies Born Better survey, we analyzed responses from 810 women who gave birth in Slovakia between March 2020 and June 2022. Via inductive content analysis we identified 4 main themes: Compassionate and Supportive Care, Medical Expertise and Care, Autonomy and Empowerment, and External Conditions. Our research has provided further evidence of the multifaceted nature of childbirth experience. The childbirth experience in a period of general adversity (such as pandemics) may have not been captured in this research or it may not significantly differ from periods of non-adversity. What is important for women remain unchanged - compassionate and supportive care from healthcare providers. Based on our findings we propose improvements in maternal healthcare during childbirth. These improvements aim not only to improve womeńs childbirth experience but also foster better healthcare outcomes for professionals and hospitals.

## Introduction

The birth experience has been referred to as a complex and multidimensional concept that expands far beyond satisfaction derived from the birth or professional care [1]. Modern perspectives emphasize the importance of a positive childbirth experience. The World Health Organization [2] defines a positive birth experience as one that meets or exceeds women’s personal and sociocultural expectations, ensuring a healthy baby is born in a clinically and psychologically safe environment with continuous support. It emphasizes that healthcare providers must offer respectful maternity care, maintaining women’s dignity, confidentiality, and privacy, and avoiding any form of mistreatment. Additionally, women should be informed about all procedures and can make free choices throughout the childbirth process.

A variety of medical and psychological factors can influence the overall childbirth experience. Birth experiences are significantly influenced by medical factors such as the type of birth, the degree of medical intervention, and the presence of skilled healthcare providers [3–6]. Psychological factors are also important in contributing to the shaping of the childbirth experience. Positive birth experiences are linked to interpersonal factors such as a respectful and trustworthy relationship with the midwife [7], empathetic, compassionate, and attentive personnel, and provision of respectful maternity care by personnel [7–13]. Positive interaction with medical personnel creates an environment perceived as supportive and safe which is a precondition for gaining control during birth [7]. Control and having one’s expectations about labor met are the leading predictors of childbirth satisfaction [5,14,15].

Positive experiences with childbearing enhance psychological accommodation for women during the postpartum period; they lead to high self-esteem, efficacy, maternal-child attachment, and satisfaction with the maternal role [15–17] while negative experience can have significant short- and long-term consequences, including mental health problems [18–21], problems in sexual functioning [22] and future reproductive decisions [23]. Negative childbirth experience can have a substantial effect not only on woman, but also on a newborn and their interaction [24,25].

In summary, research highlights that in order to achieve positive birth experience, obstetric care should be individualized, tailored to meet women’s personal and sociocultural expectations and that this care is provided in a psychologically safe environment [2,26].

As childbirth experience can have various effect on a woman and a child, it is crucial to investigate womeńs perspectives and experiences with childbirth and identify potential problems and barriers to provide adequate obstetric care. Especially in times of crises. After COVID-19 outbreak, modifications to or reductions in antenatal, perinatal, and postnatal care were globally implemented with the goal of minimizing infection risks for women, newborns, and healthcare staff alike [27]. These adjustments included decreasing the frequency of doctor’s visits, shifting to online consultations, prohibiting companions during labor or visitors in general, and separating women from their newborns [27–29]. Furthermore, maternity care during the pandemic did not meet the WHO-defined standards of quality maternity care, particularly in terms of effective communication between staff and women, respect, confidentiality, and emotional support [30,31]. These all adjustments and changes affected womeńs childbirth experience and might lead to worse experience [30–33]. However, some positive outcomes have also been reported such as shortened hospital stay, hospital safety measures supported their feeling of security the privacy and peace that resulted from the health restrictions [32–34].

Only few studies have looked at these dynamics from the perspective of women and Slovakia has not been included in this restricted number. We decided to meet this gap and try to obtain information on women’s experiences by using the Babies Born Better survey.

## Methods

Our study is grounded in the Babies Born Better survey. This long-term international survey collects data on womeńs experiences with maternity care. The goal of the project is to identify and understand what needs to be changed in an effort to improve childbirth care and then apply these findings in practice across Europe, ensuring that women and children have the best possible conditions for childbirth care [35]. Currently, the project is managed by the University of Central Lancashire (UCLan) in the United Kingdom.

### Procedure

The data we worked with were collected through an online questionnaire as part of the Babies Born Better project. The questionnaire contained 23 questions, both open and closed, focusing on sociodemographic characteristics (age, parity, standard of living), clinical factors (problems during pregnancy, type of birth), experiences with childbirth care, and a section for women’s free comments.

The questionnaire was published on various websites (Facebook, Twitter, Forums) where women and pregnant women gather. At the beginning of the questionnaire, women were instructed to fill it out only if they had given birth in Slovakia in the last three years from the date they were completing the questionnaire.

### Research sample

A total of 1,186 women completed the questionnaire during the survey period which was open from June 2020 to July 2022. After data cleaning, the final research sample comprised 810 participants who gave birth in healthcare facilities across Slovakia between March 2020 and June 2022. The first participant submitted their response on August 3, 2020, and the last entry was recorded on July 18, 2022. Upon analyzing the collected data, we found that the age of the participants ranged from 18 to 42 years, with an average age of 29.7. Detailed information on the sample is stated in Table 1 and 2.

**Table 1.**
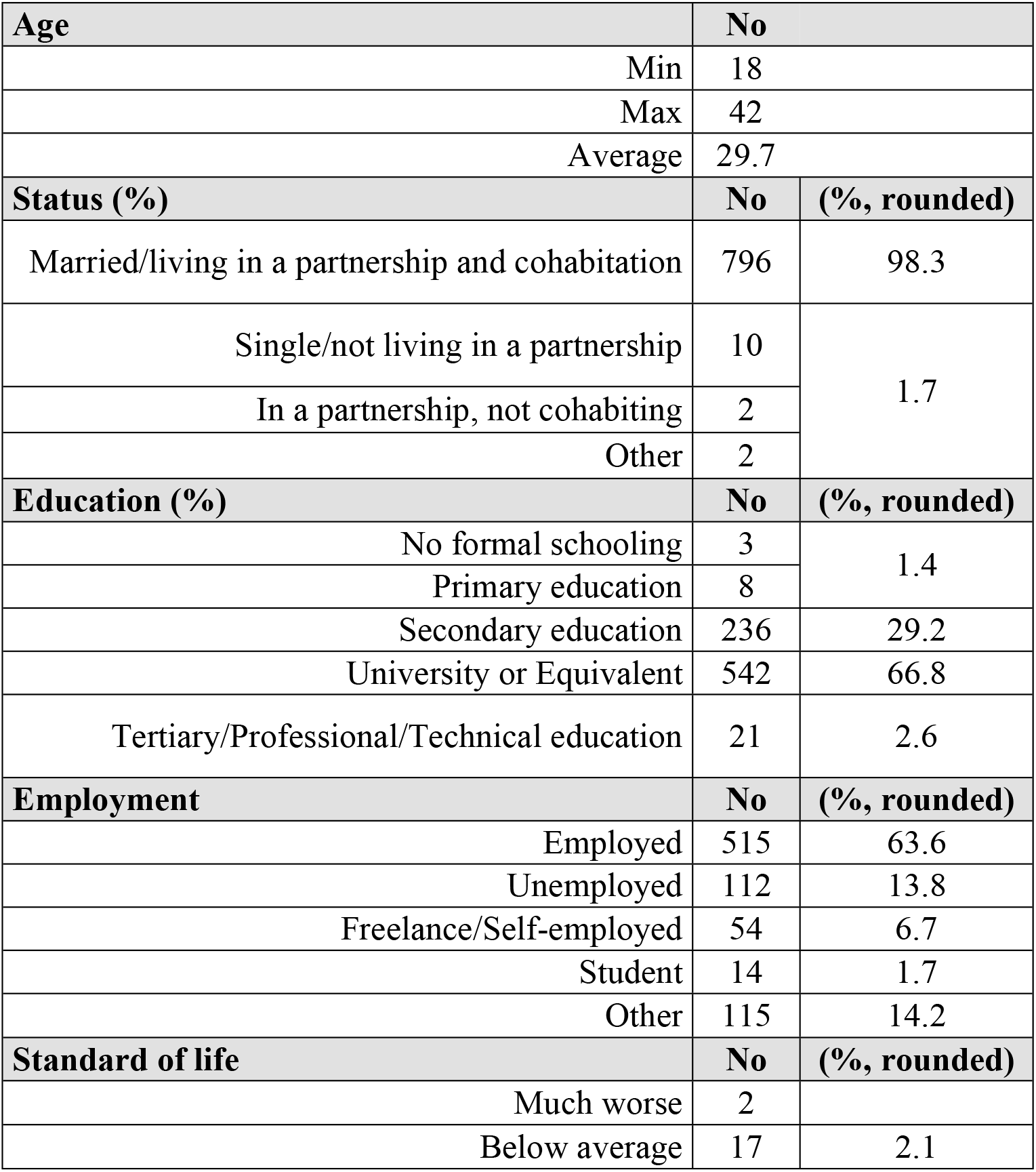
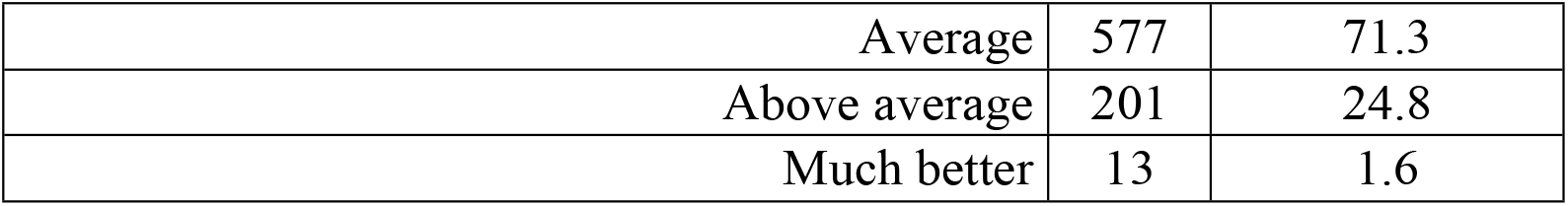
Demographics information of the sample.

**Table 2.** Childbirth indicators of the sample.

| **Weeks of pregnancy** | **No** | **(%, rounded)** |
| --- | --- | --- |
| Up to 37. week | 73 | 9.0 |
| 38. to 41 week | 666 | 82.2 |
| More than 42. week | 70 | 8.6 |
| Empty | 1 |  |
| **Pregnancy** | **No** | **(%, rounded)** |
| Without problems | 441 | 54.5 |
| With problems | 369 | 45.5 |
| **Problems in pregnancy** | **No** | **(%, rounded)** |
| Minor medical problems | 257 | 31.7 |
| Minor non-medical problems | 78 | 9.6 |
| Severe medical problems | 40 | 4.9 |
| Severe non-medical problems | 9 | 1.1 |
| **Type of birth** | **No** | **(%, rounded)** |
| Normally with no assistance | 550 | 67.9 |
| By a caesarean section, due to an emergency in labour | 131 | 16.2 |
| By a caesarean section, planned in pregnancy | 94 | 11.6 |
| With the help of ventouse (suction) or forceps or other | 35 | 4.3 |

### Ethical considerations

This survey underwent an ethical approval process at the University of Central Lancashire, UK, Committee for Ethics and Integrity (STEMH 449 Amendment_1Jun20). All personal data, whether in digital or paper form, will be stored and processed in accordance with the UK Data Protection Act (1998) as well as in line with the guidelines of the University of Central Lancashire, UK. All participants were over eighteen years old and provided written informed consent prior to beginning the survey. The survey was fully anonymous, and participants were informed that once they submitted their responses, data withdrawal would not be possible.

### Data analysis

We worked with data collected as part of the Babies Born Better project. Closed questions (sociodemographic data and information about pregnancy and childbirth) were analyzed using the Excel program Microsoft Office Excel 2023 v2302.

We analysed open questions regarding appreciations (“*In the place where you gave birth, what were the three most positive experiences of your care?*”) and suggestions for improvements (“*What do you think could have made your experience better?*”). We conducted a qualitative analysis using qualitative content analysis techniques. Inductive category formation is a fundamental part of this process, involving the creation of categories directly from the data in a bottom-up or data-driven manner [36,37]. Our process involved the following steps: (1) reading the entire data set to gain an overview; (2) coding each response; (3) refining and grouping initial codes into broader themes and finally (4) comparing and re-evaluating codes and themes [36,37]. Each author independently followed this procedure, and we discussed and resolved any discrepancies in themes until we reached a consensus.

Unlike deductive approaches, inductive category formation does not depend on pre-existing theories. Instead, it allows patterns and themes to emerge naturally from the data. This flexibility helps uncover new insights and a deeper understanding of the phenomenon being studied. The categories that result from this process are then organized into a coherent coding scheme, which can be used for further analysis [36]. Additionally, we incorporated quantitative analysis to enhance the rigor of our qualitative content analysis by including measurable elements, such as frequency counts of codes and themes, within the data set. These elements can reveal significant trends and relationships [37]. This dual approach allows researchers to combine the in-depth understanding of qualitative data with the statistical robustness of quantitative methods, leading to more reliable and valid conclusions.

## Results

The inductive content analysis revealed a structured hierarchy of categories and themes. (Table 3) Below are the detailed findings, highlighting the frequencies of each category and theme (Table 4). Regarding most positive experiences, 2,514 answers were identified. 2,472 answers were identified for suggestions. All answers were then gathered into 27 categories and subsequently grouped into 4 main themes plus uncategorized answers and no answers.

**Table 3.**
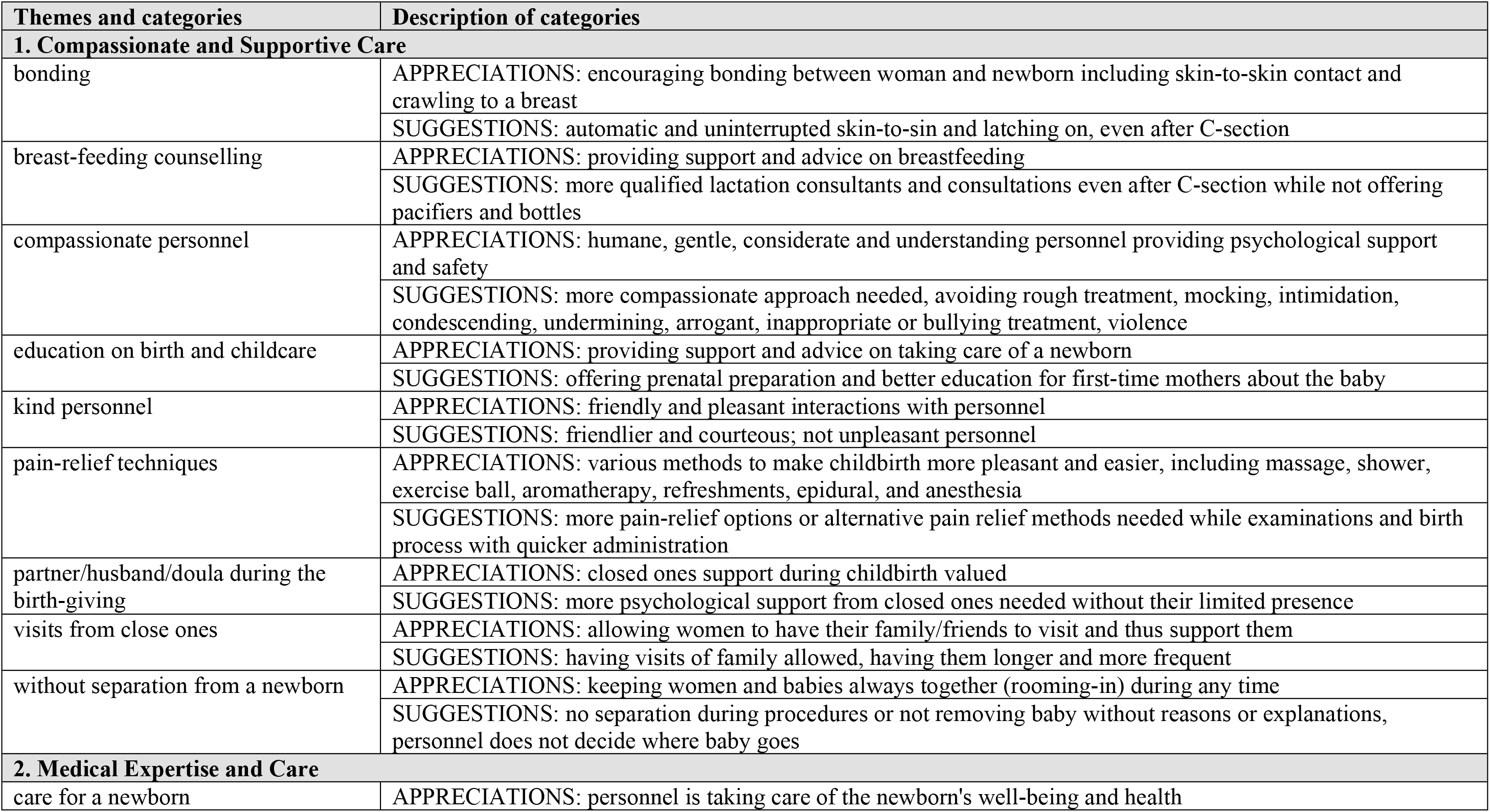
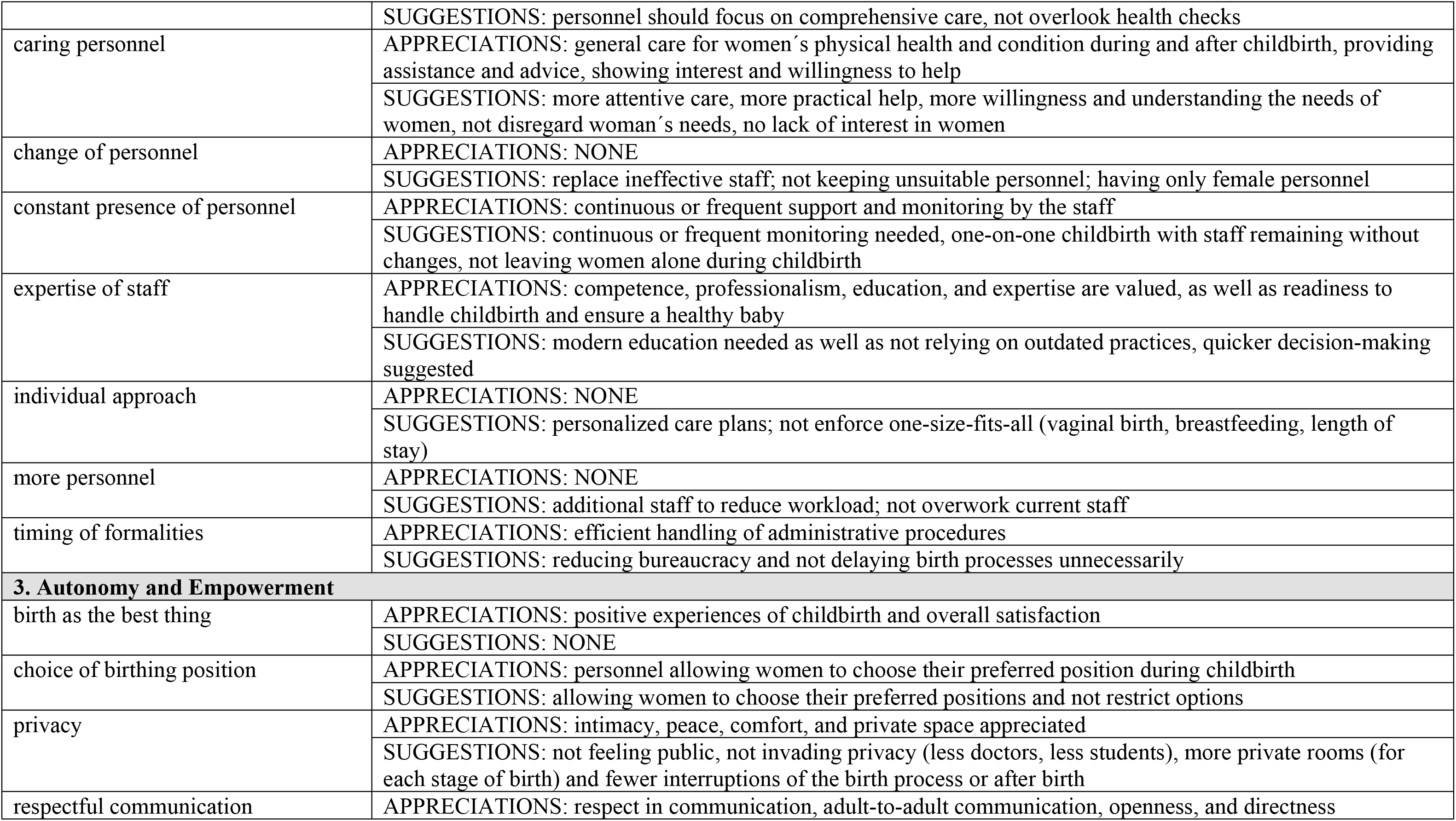
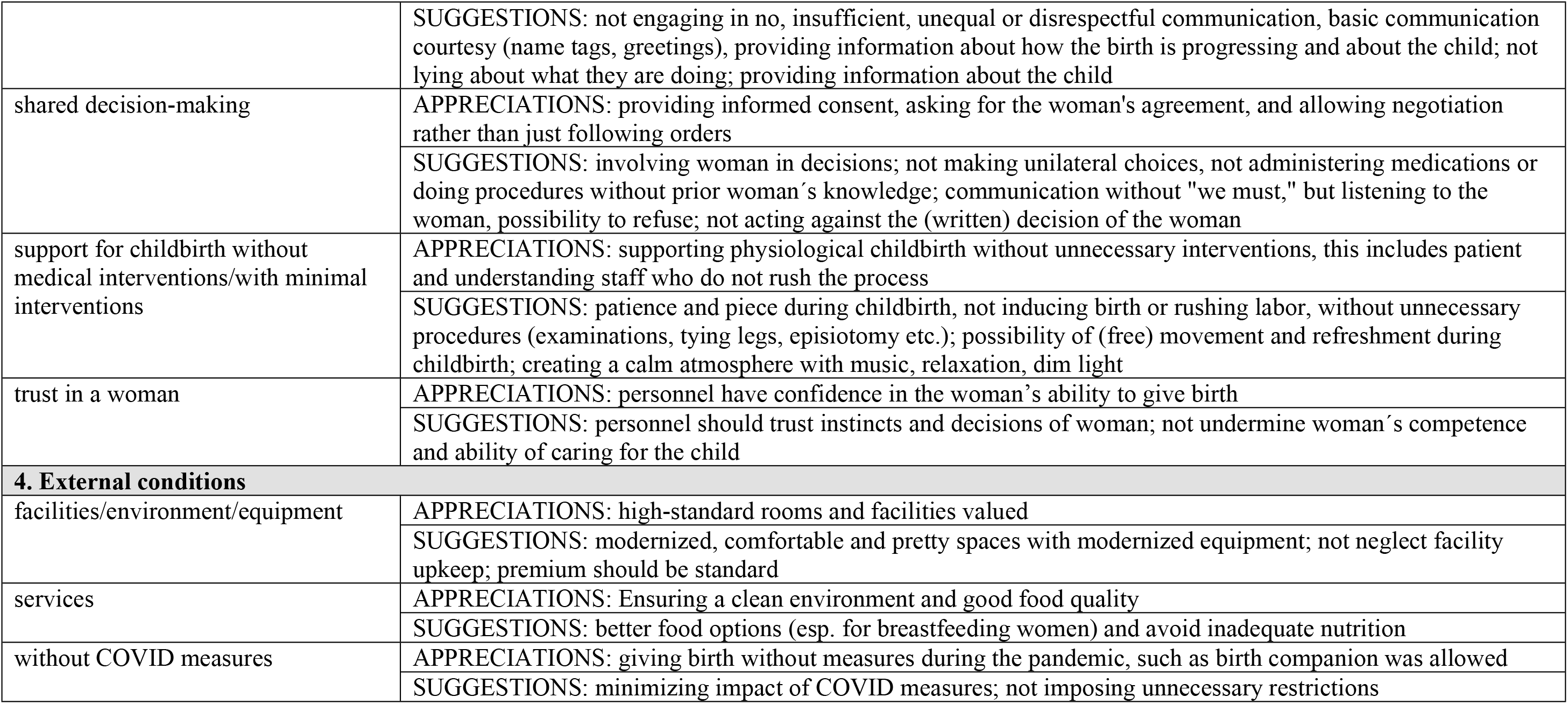
Themes and categories with descriptions in the final coding framework.

| Themes and categories | Description of categories |
| --- | --- |
| **1. Compassionate and Supportive Care** |  |
| bonding | APPRECIATIONS: encouraging bonding between woman and newborn including skin-to-skin contact and crawling to a breast |
|  | SUGGESTIONS: automatic and uninterrupted skin-to-sin and latching on, even after C-section |
| breast-feeding counselling | APPRECIATIONS: providing support and advice on breastfeeding |
|  | SUGGESTIONS: more qualified lactation consultants and consultations even after C-section while not offering pacifiers and bottles |
| compassionate personnel | APPRECIATIONS: humane, gentle, considerate and understanding personnel providing psychological support and safety |
|  | SUGGESTIONS: more compassionate approach needed, avoiding rough treatment, mocking, intimidation, condescending, undermining, arrogant, inappropriate or bullying treatment, violence |
| education on birth and childcare | APPRECIATIONS: providing support and advice on taking care of a newborn |
|  | SUGGESTIONS: offering prenatal preparation and better education for first-time mothers about the baby |
| kind personnel | APPRECIATIONS: friendly and pleasant interactions with personnel |
|  | SUGGESTIONS: friendlier and courteous; not unpleasant personnel |
| pain-relief techniques | APPRECIATIONS: various methods to make childbirth more pleasant and easier, including massage, shower, exercise ball, aromatherapy, refreshments, epidural, and anesthesia |
|  | SUGGESTIONS: more pain-relief options or alternative pain relief methods needed while examinations and birth process with quicker administration |
| partner/husband/doula during the birth-giving | APPRECIATIONS: closed ones support during childbirth valued |
|  | SUGGESTIONS: more psychological support from closed ones needed without their limited presence |
| visits from close ones | APPRECIATIONS: allowing women to have their family/friends to visit and thus support them |
|  | SUGGESTIONS: having visits of family allowed, having them longer and more frequent |
| without separation from a newborn | APPRECIATIONS: keeping women and babies always together (rooming-in) during any time |
|  | SUGGESTIONS: no separation during procedures or not removing baby without reasons or explanations, personnel does not decide where baby goes |
| **2. Medical Expertise and Care** |  |
| care for a newborn | APPRECIATIONS: personnel is taking care of the newborn's well-being and health |
|  | SUGGESTIONS: personnel should focus on comprehensive care, not overlook health checks |
| caring personnel | APPRECIATIONS: general care for women's physical health and condition during and after childbirth, providing assistance and advice, showing interest and willingness to help |
|  | SUGGESTIONS: more attentive care, more practical help, more willingness and understanding the needs of women, not disregard woman's needs, no lack of interest in women |
| change of personnel | APPRECIATIONS: NONE |
|  | SUGGESTIONS: replace ineffective staff; not keeping unsuitable personnel; having only female personnel |
| constant presence of personnel | APPRECIATIONS: continuous or frequent support and monitoring by the staff |
|  | SUGGESTIONS: continuous or frequent monitoring needed, one-on-one childbirth with staff remaining without changes, not leaving women alone during childbirth |
| expertise of staff | APPRECIATIONS: competence, professionalism, education, and expertise are valued, as well as readiness to handle childbirth and ensure a healthy baby |
|  | SUGGESTIONS: modern education needed as well as not relying on outdated practices, quicker decision-making suggested |
| individual approach | APPRECIATIONS: NONE |
|  | SUGGESTIONS: personalized care plans; not enforce one-size-fits-all (vaginal birth, breastfeeding, length of stay) |
| more personnel | APPRECIATIONS: NONE |
|  | SUGGESTIONS: additional staff to reduce workload; not overwork current staff |
| timing of formalities | APPRECIATIONS: efficient handling of administrative procedures |
|  | SUGGESTIONS: reducing bureaucracy and not delaying birth processes unnecessarily |
| **3. Autonomy and Empowerment** |  |
| birth as the best thing | APPRECIATIONS: positive experiences of childbirth and overall satisfaction |
|  | SUGGESTIONS: NONE |
| choice of birthing position | APPRECIATIONS: personnel allowing women to choose their preferred position during childbirth |
|  | SUGGESTIONS: allowing women to choose their preferred positions and not restrict options |
| privacy | APPRECIATIONS: intimacy, peace, comfort, and private space appreciated |
|  | SUGGESTIONS: not feeling public, not invading privacy (less doctors, less students), more private rooms (for each stage of birth) and fewer interruptions of the birth process or after birth |
| respectful communication | APPRECIATIONS: respect in communication, adult-to-adult communication, openness, and directness |
|  | SUGGESTIONS: not engaging in no, insufficient, unequal or disrespectful communication, basic communication courtesy (name tags, greetings), providing information about how the birth is progressing and about the child; not lying about what they are doing; providing information about the child |
| shared decision-making | APPRECIATIONS: providing informed consent, asking for the woman's agreement, and allowing negotiation rather than just following orders |
|  | SUGGESTIONS: involving woman in decisions; not making unilateral choices, not administering medications or doing procedures without prior woman's knowledge; communication without "we must," but listening to the woman, possibility to refuse; not acting against the (written) decision of the woman |
| support for childbirth without medical interventions/with minimal interventions | APPRECIATIONS: supporting physiological childbirth without unnecessary interventions, this includes patient and understanding staff who do not rush the process |
|  | SUGGESTIONS: patience and piece during childbirth, not inducing birth or rushing labor, without unnecessary procedures (examinations, tying legs, episiotomy etc.); possibility of (free) movement and refreshment during childbirth; creating a calm atmosphere with music, relaxation, dim light |
| trust in a woman | APPRECIATIONS: personnel have confidence in the woman's ability to give birth |
|  | SUGGESTIONS: personnel should trust instincts and decisions of woman; not undermine woman's competence and ability of caring for the child |
| **4. External conditions** |  |
| facilities/environment/equipment | APPRECIATIONS: high-standard rooms and facilities valued |
|  | SUGGESTIONS: modernized, comfortable and pretty spaces with modernized equipment; not neglect facility upkeep; premium should be standard |
| services | APPRECIATIONS: Ensuring a clean environment and good food quality |
|  | SUGGESTIONS: better food options (esp. for breastfeeding women) and avoid inadequate nutrition |
| without COVID measures | APPRECIATIONS: giving birth without measures during the pandemic, such as birth companion was allowed |
|  | SUGGESTIONS: minimizing impact of COVID measures; not imposing unnecessary restrictions |

**Table 4.** Themes and categories and their frequencies.

| Themes and categories | Appreciations<br>(N) | Appreciations<br>(% theme,<br>rounded) | Appreciations<br>(% total,<br>rounded) | Suggestions<br>(N) | Suggestions<br>(% theme,<br>rounded) | Suggestions<br>(% total,<br>rounded) |
| --- | --- | --- | --- | --- | --- | --- |
| **1. Compassionate and Supportive Care** | **753** |  | **30.0** | **429** |  | **17.4** |
| bonding | 120 | 15.9 | 4.8 | 63 | 14.7 | 2.6 |
| breast-feeding counselling | 16 | 2.1 | 0.6 | 62 | 14.5 | 2.5 |
| compassionate personnel | 177 | 23.5 | 7.0 | 69 | 16.1 | 2.8 |
| education on birth and childcare | 1 |  |  | 11 | 2.6 | 0.5 |
| kind personnel | 267 | 35.5 | 10.7 | 90 | 21.0 | 3.6 |
| pain-relief techniques | 71 | 9.4 | 2.8 | 29 | 6.8 | 1.2 |
| partner/husband/doula during the birth-giving | 90 | 12.0 | 3.6 | 65 | 15.2 | 2.6 |
| visits from close ones | 1 |  |  | 8 | 1.9 | 0.3 |
| without separation from a newborn | 10 | 1.3 | 0.4 | 32 | 7.5 | 1.3 |
| **2. Medical Expertise and Care** | **462** |  | **18.4** | **169** |  | **6.8** |
| care for a newborn | 32 | 6.9 | 1.3 | 10 | 5.9 | 0.4 |
| caring personnel | 237 | 51.3 | 9.4 | 63 | 37.3 | 2.6 |
| change of personnel | - | - | - | 10 | 5.9 | 0.4 |
| constant presence of personnel | 8 | 1.7 | 0.3 | 11 | 6.5 | 0.5 |
| expertise of staff | 184 | 39.8 | 7.3 | 41 | 24.3 | 1.7 |
| individual approach | - | - | - | 14 | 8.3 | 0.6 |
| more personnel | - | - | - | 8 | 4.7 | 0.3 |
| timing of formalities | 1 |  |  | 12 | 7.1 | 0.5 |
| **3. Autonomy and Empowerment** | **394** |  | **15.7** | **345** |  | **14.0** |
| birth as the best thing | 13 | 3.3 | 0.5 | - | - | - |
| choice of birthing position | 26 | 6.6 | 1.0 | 53 | 15.4 | 2.1 |
| privacy | 68 | 17.3 | 2.7 | 55 | 15.9 | 2.2 |
| respectful communication | 147 | 37.3 | 5.9 | 99 | 28.7 | 4.0 |
| shared decision-making | 64 | 16.2 | 2.6 | 36 | 10.4 | 1.5 |
| support for childbirth without medical interventions/with minimal interventions | 75 | 19.0 | 3.0 | 87 | 25.2 | 3.5 |
| trust in a woman | 1 |  |  | 15 | 4.4 | 0.6 |
| **4. External conditions** | **65** |  | **2.6** | **177** |  | **7.2** |
| facilities/environment/equipment | 53 | 81.5 | 2.1 | 124 | 70.1 | 5.0 |
| services | 11 | 16.9 | 0.4 | 45 | 25.4 | 1.8 |
| without COVID measures | 1 | 1.5 |  | 8 | 4.5 | 0.3 |
| **5. Uncategorized** | **9** |  | **0.4** | **4** |  | **0.2** |
| **6. “Nothing” as response** | **831** |  | **33.1** | **1,348** |  | **54.5** |
| **ALTOGETHER** | **2,514** |  |  | **2,472** |  |  |

## COMPASSIONATE AND SUPPORTIVE CARE

The theme of *Compassionate and Supportive Care* received significant attention, with 753 mentions of positive experiences (30 %) and 429 suggestions (17.4 %). 9 categories were included in this theme depicting personnel behaviour and procedures at the department that support women psychologically.

Regarding personnel behaviour, women highly appreciated the *kind personnel*, which was mentioned 267 times (10.7 %), highlighting the importance of friendly, pleasant, and excellent interactions with staff. In contrast, the need for kinder, more respectful, and courteous behavior was suggested 90 times (3.6 %). *Compassionate personnel* were praised 177 times (7.0 %) for being humane, gentle, considerate, and understanding, providing psychological support and safety and reducing stress. However, 69 suggestions (2.8 %) called for more compassionate approaches, avoiding rough treatment, disparagement, and inappropriate behavior.

Regarding procedures that are at the department, there were several procedures mentioned. Allowance of *Bonding* between woman and newborn, as the third most numbered code in this theme, was encouraged 120 times (4.8 %), with 63 suggestions (2.6 %) advocating for uninterrupted skin-to-skin contact even after a C-section. The presence of a *partner, husband, or doula during childbirth* was appreciated 90 times (3.6 %), while 65 suggestions (2.6 %) called for more support people. *Pain-relief techniques* were positively noted 71 times (2.8 %) for various methods, including massage, shower, exercise ball, aromatherapy, and epidurals. Suggestions for improvements in pain-relief techniques were mentioned 29 times (1.2 %), advocating for quicker administration of epidural and alternative pain management options.

In the last few codes suggestions prevailed over appreciations. *Breast-feeding counselling* was mentioned positively only 16 times (0.6 %), with 62 suggestions (2.5%) for more qualified lactation consultants. *Education on birth and childcare* was appreciated once, with 11 suggestions (0.5 %) for better prenatal preparation and postnatal education of women. *Visits from close ones* were mentioned once positively, with 8 suggestions (0.3 %) for allowing longer and more frequent visits. *Without separation from a newborn* was positively noted 10 times (0.4 %), but with 32 suggestions (1.3 %) advocating for keeping woman and baby together.

## MEDICAL EXPERTISE AND CARE

*Medical Expertise and Care* accounted for 462 mentions of positive experiences (18.4%) and 169 suggestions (6.8 %). It focuses on physical health and well-being, emphasizing essential obstetric care practices. It includes 8 categories with three being present only in suggestions.

*Caring personnel* was highly appreciated (238 times; 9.4%). Women valued general care for physical health and condition during and after childbirth, healthcare providers assistance, and interest in their condition, However, 63 suggestions (2.6 %) called for more attentive and supportive care with higher willingness and understanding of personnel. The *expertise of personnel* was highly appreciated 184 times, (7.3%) emphasizing competence, professionalism, and readiness to handle childbirth and women noticed expertise of doctors, midwives, nurses, paediatricians, anaesthesiologists and other personnel as well. Suggestions for better expertise, modern practices and quicker decision-making were mentioned 41 times (1.7 %).

*Care for a newborn* was positively recognized in 32 times (1.3 %). 10 suggestions (0.4%) called for ensuring the newborn’s well-being by healthcare professionals. *Timing of formalities* was appreciated once, with 12 improvements (0.5 %) suggesting more efficient handling of administrative procedures and not interrupting the process of birth. *Constant presence of personnel* received 8 positive mentions (0.3 %), with 11 suggestions (0.5 %) for continuous support.

Last three codes were mentioned only in suggestions. *More personnel* were suggested 8 times (0.3 %) to reduce the workload on existing staff. *Change of personnel* received 10 suggestions (0.4 %) for replacing staff members. The need for an *individual approach* was suggested 14 times (0.6 %), advocating for adapting care to the woman’s needs.

## AUTONOMY AND EMPOWERMENT

The theme of *Autonomy and empowerment* had 394 positive mentions (15.7 %) and 345 suggestions (14.0 %). 7 categories describe personnel behaviour, processes and culture of the hospital and are centered around respecting women as active agents in childbirth.

*Respectful communication* was the most frequently appreciated category, with 147 positives (5.9 %) that highlighted the importance of respectful, equal and adult-to-adult communication. However, 99 suggestions (4.0 %) called for better communication, basic communication courtesy (introducing oneself, name tags) and providing information about the childbirth and a newborn.

*Support for childbirth without medical interventions or minimal interventions* was positively noted 75 times (3.0 %), with 87 suggestions (3.5 %) calling for minimal interventions, more physiological childbirth experiences and enough time to do labour and childbirth in womeńs own pace. *Choice of birthing position* was positively mentioned 26 times (1.0 %), with 53 suggestions (2.1 %) advocating for letting women choose their birth position. *Privacy* during childbirth was positively noted 68 times (2.7%), with 55 suggestions (2.2 %) for more private settings during examinations and childbirth and not to be disturbed by staff. *Shared decision-making* was appreciated 64 times (2.6 %), with 36 suggestions (1.5 %) calling for more involvement of women in decision-making processes and asking women for their consent.

*Trust in a woman* was mentioned once positively, with 15 suggestions (0.6 %) calling for more confidence in women’s feelings and ability to give birth*. Birth as the best thing* was only mentioned in positive experiences 13 times (0.5 %) describing positive experiences of childbirth and overall satisfaction with birth.

## OTHER RESPONSES

The last theme, *External Conditions* accounted for 65 positive mentions (2.5 %) and 178 suggestions (7.2 %). The need for better *facilities, environment, and equipment* was mentioned 53 times positively (2.1 %), with 124 suggestions (5.0 %) for improvements such as nicer rooms, modern equipment, and better amenities. *Services* (cleaning and food) received 11 positive mentions (0.4 %), with 45 suggestions (1.8 %) for better food quality to support breastfeeding. The impact of *COVID-19 measures* was not mentioned positively but had 8 suggestions (0.3 %) for consideration of how these measures affect the childbirth experience.

A notable portion of responses, categorized as *nothing*, was provided 831 times in most positive experiences, representing 33.1 % of the total for this analysis. Similarly, 1,348 times (54.5 %) women did not state any suggestions that might improve their hospital experiences.

## Discussion

The results of the inductive content analysis provide a detailed understanding of womeńs experiences during childbirth under strenuous the COVID-19 outbreak conditions. We investigated experiences of women that gave birth from March 2020 to June 2022. Women formulated positives as well as suggestions regarding obstetric care in hospitals and we collected 2,514 appreciations and 2,472 suggestions for improvement that we subsequently gathered into 27 categories and 4 main themes plus uncategorized answers and no answers.

The most frequent theme (30.0 % in satisfaction and 17.4 % in suggestions) was *Compassionate and Supportive Care*, which can be figuratively named ‘*Understand Me*’. This theme included nine categories describing either staff personal behaviour or procedures related to childbirth that the department/healthcare professionals follow (regarding pain relief, presence of support people, bonding and breastfeeding, and visits). ‘*Understand Me*’ theme emphasizes empathy and emotional support during and after childbirth. Responses often reflected the humanity, kind behavior, and empathetic approach of the staff, who understand the birth process and support women in managing it better. It involves creating a positive and nurturing environment with attentive and kind interactions from healthcare providers and having significant people around, such as a doula, partner, and family. It also includes postnatal supportive care, namely encouraging bonding, breastfeeding, newborn care, and ensuring the woman is not separated from her child. In summary, the theme ‘*Understand Me*’ encompasses behaviors and practices that reduce stress, promote emotional security, and enhance women’s psychological well-being.

Our findings on *Compassionate and Supportive Care* are corroborated by previous studies that emphasize the importance of empathy and emotional support during and after childbirth across countries [7–9,11–13]. Women welcome if there is a significant adult around them that helps them to manage their labor and childbirth, such as a trustworthy midwife, partner, doula, or other companion [7–9,38]. For women as well as babies, continuous care during labor has benefits that are clinically significant. Continuous support increased probability for vaginal birth, shortened labour time, and decreased probability of surgical birth, use of any analgesia [4,39]. Moreover, continuous support diminishes negative experiences regarding childbirth [39]. Long-term effects are shown on maternal mental health. For example, intrapartum as well as postpartum social support seems to diminish risks of postpartum depression [40].

Still quite a large proportion of women in our research perceive a lack of social support, empathy, compassion. Suggestions often reflected the desired compassion, kindness, and empathetic approach of the staff. Some women expressed the need to feel that they were not just another ‘number’ to the staff. Women even encounter open forms of aggression, such as arrogant behavior, intimidation, unpleasant remarks, and insults. Unfortunately, these findings on disrespectful behavior from healthcare providers are not surprising and are also found in surveys from other countries around the world [8,11,12,41]. For instance, in Netherlands more than 11,000 women were included in the survey and they reflected between years 2015-2020. 45% of primipara and 27% multipara experienced at least one upsetting disrespect and abuse from healthcare providers during labour and birth [41]. Disrespectful maternity care does not confine only to low-income countries, but it has been recorded in high-income countries, such as the USA too [42].

Based on the results of previous qualitative studies, inhumane behavior, intimidation, coercion, and manipulation by obstetric staff was already present in Slovakia even before the pandemic [43,44]. This problem was even stronger with the onset of the pandemic, as several studies speak of non-compliance with quality standards of maternity care during the pandemic, such as those set by the WHO [30]. The quality standards of maternity care that were affected were effective communication, respect and dignity, emotional support, and availability of competent and motivated medical staff.

Overall, compassionate and supportive care is not only a whim or wish by women. It is a crucial requirement stated by women themselves that should be a part of common obstetric care. It aligns with the WHO standards [2] and guidelines stated by the International Federation of Gynecology and Obstetrics [45] as it focuses on womeńs well-being and mental health that are particularly important during and after childbirth. It has been also proven via research that it helps women to deal with birth and to articulate birth as a positive experience. Our research shows that women are sensitive to social interaction with hospital personnel and their behavior. They strive for compassionate and sensitive behavior from the personnel also in the time of adversity. The theme of *Medical Expertise and Care*, which we named ‘*Help Me*’, was the second most appreciated theme (18.4 %) and the third theme with regard to suggestions (6.8%). Unlike the ‘*Understand Me*’ theme that focuses on mental health and well-being, ‘*Help Me*’ centers on physical health and well-being. The guiding principle is the Hippocratic Oath, particularly the tenets of *first, do no harm* (non-maleficence) and beneficence. The primary goal is to ensure a safe childbirth and results in a physically healthy baby and woman.

Our findings indicate that women greatly appreciate the professionalism and dedicated care provided by healthcare staff. However, despite the positive feedback, many women suggested that there needs to be more attentive and practical care, greater interest, and willingness to assist women by healthcare personnel. Other suggestions included the adoption of modern practices, improved expertise among personnel, efficient handling of formalities, individualized approaches, and increased staffing or changes in personnel. Importance of professionalism in womeńs subjective childbirth experiences is also articulated in previous studies before pandemic [11,46–48].

In summary, the ‘*Help Me*’ theme highlights the critical role of knowledge and skills in obstetrics, and it underscores the importance of a safe and professionally supported childbirth, where the physical well-being of both woman and baby is paramount. This is once again a part of personnel factor (knowledge, skills, and behavior) that was important for women even before the pandemic. It stays important even in the time of the adversity.

Another significant theme identified in our study is ‘*Autonomy and Empowerment*’, which encompasses 7 categories with 394 (15.7 %) appreciations and 345 (14.0 %) suggestions. The theme encompasses personnel behaviour, processes and culture of the hospital that encourage women to be active participants and engaged in the process of birth. Women need to actively participate in decision-making about their childbirth and individual procedures. They highly value equitable communication, which is open, direct, and informative. Encouragement and support for physiological childbirth and related techniques are also central to this theme. Women require oversight from professionals during childbirth, but this oversight should not compromise their control, autonomy, and competence. Instead, it should foster an autonomous, self-managing woman who experiences a positive birth. Therefore, we named this theme ‘*Empower me*’.

The focus on autonomy and empowerment aligns with the World Health Organizatiońs new approaches to health literacy [49]. The goal is to foster self-efficacy in patients, in our case, women, emphasizing their autonomy and self-management. Psychological theories also underpin the importance of autonomy and empowerment. For example, self-determination theory [50] emphasizes three basic psychological needs, namely autonomy, competence, and relatedness. Recently, this theory was applied as a new framework to understand childbirth [51].

Our findings on autonomy and empowerment are corroborated by previous research too. For instance, personal control strongly predicted childbirth satisfaction [15]. In their systematic qualitative review Downe and colleagues [52] also found that women appreciate personal achievement and control thanks to shared decision making. Shared-decision making is also underscored in another systematic review [53]. Feelings of empowerment because of being able to give birth and trust in onés ability is highlighted in research by Karlström and colleagues [7] in which they analysed a very positive childbirth experience. Finally, in their discussion paper, Leinweber and colleagues [54] also emphasize feelings of being in control and being respected as parts of positive childbirth. In summary, autonomy and empowerment are parts of childbirth experience that are important for women in all periods of pandemics and non-pandemics.

While the previous themes described psychological and medical factors on the childbirth experience, the theme ‘*External conditions*’ focuses more on environment in which childbirth happens. This includes experiences with birthing spaces, rooms, and hospital facilities, including services such as food services and cleaning services. Measure regarding COVID-19 were also included here, such as wearing the masks.

In comparison with other themes, this theme seems to be undervalued as it received less than 3 or 8 % in appreciations and suggestions respectively. However, even smaller numbers are important in exploring insights. Importance of hospital environment was also highlighted in Croatian survey [11] and in Austrian as well [46].

In our research, only small portion of women explicitly commented on pandemic measures (9 altogether). One woman was glad that there was a birth companion allowed during the pandemic. On the other hand, there were suggestions to lessen restrictions regarding wearing masks and birth companion. However, this does not mean that women did not reflect on changes in maternity care. They simply did not directly label these changes in their statements. Many categories that we created could have been, and certainly were affected by the COVID-19 pandemic [55]. For example, in the theme of *medical expertise and care*, this could have been reflected in the category of *’caring staff*,’ where women may have mentioned how they appreciated the care provided by the staff but did not further explain whether this was because of staff shortages due to the pandemic, and how the staff tried to compensate for this as it was mentioned in other study [31]. In the theme of *compassionate and supportive care*, several categories could have been affected, such as *kind personnel, compassionate personnel, bonding, no separation from the newborn,* supporting people *(partner/husband/doula during childbirth), breastfeeding support, or visits from close ones*.

### Practical implications

There are several suggestions that might improve womeńs experience with childbirth not only during adversities. We can gather them into three areas: healthcare professionals and their soft skills, mental health services for healthcare professionals and, finally, willingness to change established processes of healthcare to improve women childbirth experiences. Thus, these suggestions need to have a managerial support to implement them.

First of all, healthcare professionals must possess strong communication skills. Effective communication involves providing information, explanations, and guidance, which is crucial for addressing both the physical and emotional needs of birthing women. Key characteristics of women’s experiences of safe childbirth included the need to be informed and involved through sharing and receiving trustworthy information. Conversely, experiences of unsafe childbirth were marked by a lack of meaningful information, leading to feelings of being misled or ignored [56]. Improving communication skills is thus crucial for enhancing empathy and building better women- provider relationships [57,58]. Equal communication invites women to conversation and gives them their voice back.

As the most frequent theme or category was compassion and supportive care- compassionate and kind personnel, we assume that empathetic and compassionate behavior is needed for healthcare professionals. Implementations of structured training focused on developing empathy seem to have impact on healthcare providers and their empathetic behavior [59,60]. Furthermore, we need to take into account that relationship between healthcare providers and women is crucial in providing healthcare. Taking the time to get to know women as individuals and understanding their unique needs and concerns can foster a more empathetic approach. This relationship-centered care is a personalized care that enhances and promotes empathy [61]. Patient engagement in care decisions leads to better health outcomes and higher patient satisfaction [62].

Moreover, we need to take care of healthcare professionals too, as they are often forgotten. Ensuring and promoting positive childbirth experience can be very exhausting and lead to decrease of compassion satisfaction and increase in compassion fatigue. Therefore, taking care of mental health of professionals is one of the key elements in keeping the high standard of healthcare. We can encourage healthcare providers to engage in reflective writing about their experiences with women which can help them process emotions and develop a deeper understanding of womeńs perspectives. Reflective writing has been shown to enhance empathy and improve patient care [63,64]. Moreover, also promoting mindfulness practices and self-care among healthcare staff can reduce burnout and increase their capacity for empathy. As has been shown, mindfulness training helps healthcare providers manage stress, burnout and improve empathy and patient-centered care [65].

And finally, we also need to create atmosphere at hospitals that is open to feedback and changes. A large portion of responses labelled as "nothing" appeared frequently among positive experiences, and similarly, many respondents offered no suggestions for improving their hospital experience. These responses indicate that a significant number of women did not have specific positive feedback or experiences to report nor they can identify possible gaps for improvements. We might also assume that they might be disinterested, not engaged in their childbirth process so much. We also might assume that there is specific culture in Slovakian hospitals. Maternity care in Slovakia is institutionalized and monopolized, provided solely in hospitals and largely shaped by male obstetricians [55]. The dominance of male obstetricians also influences power dynamics in the field, often disadvantaging pregnant and birthing women [55]. However, regularly seeking and reflecting on womeńs feedback can help healthcare providers understand the impact of their behavior and make necessary adjustments to improve empathetic care. Patient feedback is a valuable tool for improving quality in healthcare [66].

### Limits

Despite the interesting results, our study has several limitations. We consider the online data collection method a limitation, as it may have resulted in insufficient control over participants and the accuracy of the data, as well as the inability to clarify potential ambiguities. It is possible that individuals who did not meet the criteria for the research sample, such as women who had not given birth at all or those who gave birth in another country, might have completed the questionnaire. Some women were not included in the research simply because they did not have a computer, internet access, or the necessary skills to participate in online research. Additionally, some questions might have been misinterpreted by participants, affecting the relevance of the responses. Consequently, we had to remove responses from several participants during data cleaning because the responses were unclear, resulting in a loss of some data.

Another limitation concerns the sample characteristics. The online survey was not aimed at obtaining a representative sample, and we were aware of this. Nevertheless, we compared the sample characteristics with data from 2022 published by the Slovak National Health Information Centre [67]. Our sample was partly biased because the majority had a university education (66.8 % in our sample vs. 31.7 % in national sample) and were living in a relationship or married (98.3 in our sample vs 58.4 % in national sample). However, age (most of the women were from age range 30-34- 38.1% in our sample vs. 31.0 % in national sample) and type of birth (67.9 % vaginal and 32.1 % operative birth in our sample vs. 67.1 % vaginal and 32.9 % operative birth in national sample) were similar to the data provided by the Slovak National Health Information Centre.

The design of the questionnaire, with several open-ended questions, can also be considered a limitation, as participants might have been unwilling to fill them out, leading them to skip these questions. This could be why open-ended questions often remained unanswered. A lot of answers were a single-word answer which might also create a bias since it was our interpretation of the words based on which we analysed the data.

## Conclusion

Our research has provided further evidence of the multifaced nature of childbirth experience. Our findings support a combination of four factors in forming childbirth experience, namely compassionate and supportive care, medical expertise and care, autonomy and empowerment of women and external conditions. These findings align with other studies conducted both before and after the COVID-19 outbreak. In general, women overwhelmingly prefer compassionate and supportive care from healthcare providers. They desire personnel who understand them and exhibit kind and empathetic behaviour that can psychologically support them during and after their childbirth. In practical terms, it means mainly having friendly and pleasant interactions with women and showing verbal and non-verbal understanding.

Our research indicates that if there has been a change in the childbirth experience, we were unable to capture it with this survey. We noticed answers regarding COVID measure (wearing masks, limitation of a companion during and after the birth, limited or shortened visits of closed ones, separation from a newborn) that had been explicitly stated in previous research as being the results of the COVID measures. However, they were not so frequently present in our survey, as one would have expected. Also, these situations (such as limitation of a companion during and after the birth, limited or shortened visits of closed ones, separation from a newborn) may have been present even before the pandemics. However, the most relevant aspect of womeńs childbirth experience seems to remain unchanged - compassionate and supportive behavior from personnel.

Based on our findings we propose improvements in maternal healthcare mainly during childbirth. These improvements aim not only to improve womeńs childbirth experience but also foster better healthcare outcomes for professionals and hospitals.

## Data Availability

Data are available from the corresponding author with the permission of Babies Born Better Collaborative Group.

## Acknowledgements

We would like to thank the participants for their participation in this study.

This paper is based on the Babies Born Better project, initiated as part of COST Action IS0907 and further developed under EU COST Action IS1405: BIRTH (“Building In Optimising Childbirth Intrapartum Research Through Health” - an interdisciplinary, whole-system approach to understanding and contextualizing physiological labor and birth) supported by the COST (European Cooperation in Science and Technology) program within EU Horizon 2020. We acknowledge the contributions of everyone involved in developing and conducting the Babies Born Better Survey. Further information about the project, Steering Group, and Country Coordinators can be found at: http://www.babiesbornbetter.org/about/.

## Author Contributions

Conceptualization: Greškovičová Investigation: Greškovičová Data Analysis: Greškovičová, Němcová, Šiková

Writing – original draft: Greškovičová, Němcová, Šiková Writing – review & editing: Greškovičová, Němcová, Šiková

